# The spatial percept of tinnitus is associated with hearing asymmetry: subgroup comparisons

**DOI:** 10.1101/2020.05.05.20073999

**Authors:** Eleni Genitsaridi, Theodore Kypraios, Niklas K. Edvall, Natalia Trpchevska, Barbara Canlon, Derek J. Hoare, Christopher R. Cederroth, Deborah A. Hall

## Abstract

The spatial percept of tinnitus is hypothesized as an important variable for tinnitus subtyping. Hearing asymmetry often associates with tinnitus laterality, but not always. One of the methodological limitations for cross-study comparisons is how the variables for hearing asymmetry and tinnitus spatial perception are defined. In this study, data from two independent datasets were combined (n= 833 adults, age ranging from 20 to 91 years, 404 males, 429 females) to investigate characteristics of subgroups with different tinnitus spatial perception focusing on hearing asymmetry. Three principle findings emerged. First, a hearing asymmetry variable emphasizing the maximum interaural difference most strongly discriminated unilateral from bilateral tinnitus. Merging lateralized bilateral tinnitus (perceived in both ears but worse in one side) with unilateral tinnitus weakened this relationship. Second, there was an association between unilateral tinnitus and ipsilateral asymmetric hearing. Third, unilateral and bilateral tinnitus were phenotypically distinct, with unilateral tinnitus being characterized by older age, asymmetric hearing, more often wearing one hearing aid, older age at tinnitus onset, shorter tinnitus duration, and higher percentage of time being annoyed by tinnitus. We recommend that careful consideration is given to the definitions of hearing asymmetry and tinnitus spatial perception in order to improve the comparability of findings across studies.

## Introduction

There is emerging evidence indicating that tinnitus percepts with different spatial profiles might represent subtypes with different mechanisms (Maas et al., 2017, Vanneste et al., 2011, Cuny et al., 2004). In addition, it has been shown that tinnitus laterality tends to associate with hearing asymmetry (Cahani et al., 1984, Tsai et al., 2012), however, this is not always the case (Lee et al., 2019).

One of the methodological limitations for cross-study comparisons is how hearing asymmetry and tinnitus spatial perception are operationally defined. There is no single established method for defining asymmetric hearing. Asymmetry can be based on the average interaural difference (ID) of specific audiometric frequencies or a frequency range, the value of the maximum difference in one or more frequencies, or a combination of characteristics. Many different approaches have been documented (Cahani et al., 1984, Caldera and Pearson, 2000, Cheng and Wareing, 2012, Hendrix et al., 1990, Hojjat et al., 2017, Jeffery et al., 2016, Mangham, 1991, Margolis and Saly, 2008, National Guideline Centre UK, 2018, Tsai et al., 2012, Urben et al., 1999). Examples from clinical practice also differ. In the UK, the British Academy of Audiology considers a diagnosis of asymmetric hearing when there is an interaural difference of *20 dB* or more in at least two consecutive frequencies at 0.5, 1, 2, 4 and 8 kHz (Jeffery et al., 2016). However, also in the UK, the National Institute for Health and Care Excellence (NICE) recommendation considers an onward referral for Magnetic Resonance Imaging (MRI) when there is an interaural difference of *15 dB* or more in two consecutive frequencies at 0.5, 1, 2, 4 and 8 kHz (National Guideline Centre UK, 2018). Based on 1490 audiograms from military personnel, Caldera and Pearson (2000) showed that the prevalence of hearing asymmetry could have a more than 100-fold variation (varying from 543 to 77,242 per 100,000) depending on the definition used for asymmetry. The task becomes even more complicated when the audiometric profile is sampled more comprehensively at mid-octave frequencies and extended high frequencies above the conventional cut-off at 8 kHz, as is often the case in research settings. One proposed solution could be to measure the area under the audiogram curve after interpolating in-between frequencies (König et al., 2006). For research purposes, some have sought to define the optimum asymmetry metric depending on the hypothesis. For example, Tsai et al. (2012) investigated how different asymmetry metrics can predict tinnitus laterality. They concluded that a maximum threshold difference averaged to the adjacent second maximum of at least 15 dB difference was the optimum predictor. However, this has not been independently verified. Examples of different definitions for hearing asymmetry reported in the literature, and their application are shown in Supplementary Table 1 and 2 respectively. Importantly, none of these measures included extended high frequency audiometric thresholds.

As in asymmetric hearing, there is no standard method for defining the spatial percept of tinnitus. Tinnitus can be perceived anywhere in space (Searchfield et al., 2015), but to localize the percept of tinnitus requires psychophysical testing procedures. Instead, studies more often rely on self-report and limit inquiry to whether tinnitus is perceived in one or both ears or in the head. Many studies use a binary classification of unilateral and bilateral tinnitus, although response options can be extended to include: in the right ear, in the left ear, in both ears equally, in both ears but worse in the right or left ear, and inside the head or elsewhere (Langguth et al., 2007, Nuttall et al., 2004). The challenge here is how to pool such response options to form characteristics that define meaningful subgroups.

Table 1 proposes four potential summary variables for tinnitus spatial perception. These discriminate the percept of tinnitus that is clearly restricted to one ear (unilateral) from that where tinnitus is perceived equally in both ears (bilateral). They also consider cases that are less distinct; where tinnitus is in both ears but more on one side than the other or is somewhere inside the head. The characterization of being lateralized or non-lateralized is used to discriminate percepts based on whether there is a dominance in one side (left or right) or not. There is some degree of subjectivity in determining whether the less distinct lateralized bilateral cases should be categorized with unilateral or bilateral tinnitus. Reasonable justifications could be made to categorize a participant who experiences tinnitus in both ears but worse on one side, either as a case of bilateral tinnitus or unilateral tinnitus.

**Table 1.**
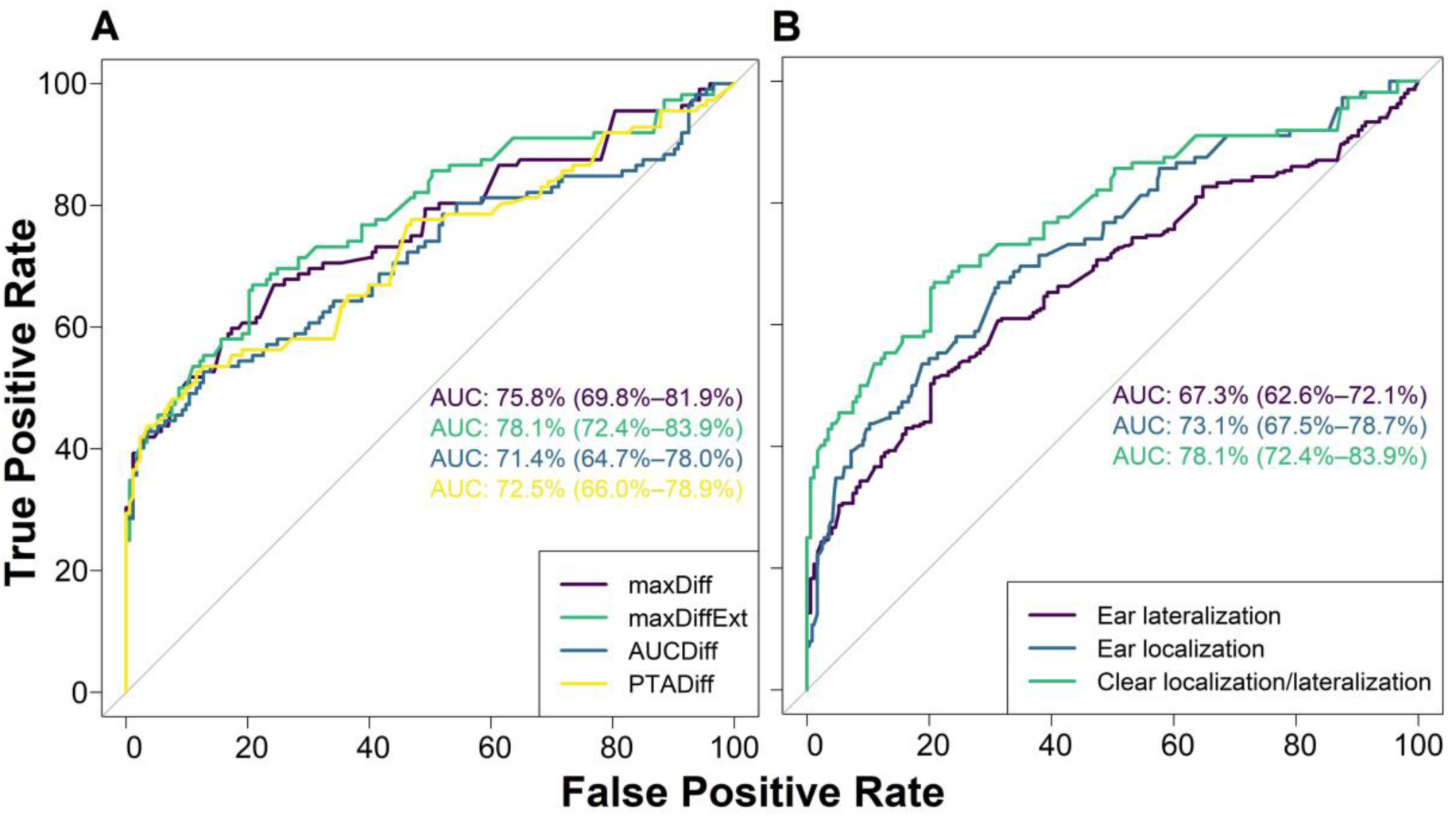
Summary labelling of response options for tinnitus spatial perception.

In this study, we combined two independent datasets to address the following research questions.

1. Which definition of hearing asymmetry reliably discriminates unilateral from bilateral tinnitus? We also explored whether participants reporting tinnitus in both ears but worse in one ear should be classified as unilateral or bilateral tinnitus cases.
2. Does the pattern of hearing asymmetry differ between tinnitus and non-tinnitus cases, and across different spatial tinnitus percepts in those reporting tinnitus?
3. What are phenotypic characteristics of subgroups with unilateral or bilateral tinnitus?

## Methods

### Dataset description

The two independent datasets were from the Swedish Tinnitus Outreach Project (STOP) Sweden and the NIHR Nottingham Biomedical Research Centre (BRC) UK. The STOP dataset analyzed was a subset from a population-based tinnitus specific database (Swedish Tinnitus Outreach Project, 2015). The BRC dataset analyzed was a collection of published data from three previous tinnitus clinical studies conducted by some of the authors (Davies et al., 2014, Hoare et al., 2012, Hoare et al., 2014). Each of these studies had received ethical approval from the National Research Ethics Committee (Nottingham or Derby, UK). For the STOP project, ethical approval was granted by the local ethics committee *“Regionala etikprövningsnämnden”* in Stockholm (2015/2129-31/1). The two datasets included a number of common variables and were composed of phenotypical information (both general and tinnitus specific) that had been collected using various hearing tests and questionnaires, including the Tinnitus Sample Case History Questionnaire (TSCHQ; Langguth et al., 2007) and the Hyperacusis Questionnaire (HQ; Khalfa et al., 2002). For the BRC dataset, pure tone audiometry was conducted manually by an examiner using a Siemens Unity 2 system and Sennheiser HDA 200 headphones. For the STOP dataset, fixed frequency Bekesy audiometry was done using the Astera 2 audiometer (Otometrics) and Sennheiser HDA 200 headphones. In both cases, frequencies from 0.125 kHz to 14 kHz were tested in sound-proofed conditions. Thresholds greater than the audiometer limit were given a standardized value of 110 dB HL. Details of all the included variables can be found in Supplementary Table 3.

Data for participants without pure tone audiometry (n=10) were excluded from further analyses. Data for participants with missing responses to the question ‘Where do you perceive your tinnitus?’ (n=19), and cases reporting tinnitus ‘elsewhere’ (n=12) were also excluded. From an initial sample of 612 tinnitus cases, this left 571 for analysis (n=382 from the STOP and n=189 from the BRC databases). Data from 262 non-tinnitus cases were also available from the STOP database. The mean age from the total sample (n=833) was 53 years, ranging from 20 to 91. There were 404 males and 429 females.

Participants with tinnitus across datasets differed significantly in terms of age, mean audiometric hearing thresholds, hearing aid use, presence of headaches and balance disorders, tinnitus duration and age at onset, spatial perception of tinnitus, stress influence on tinnitus and percentage of time being annoyed by tinnitus. This information is shown in Supplementary Table 4. These observations fall within the variability that would be expected, considering the differences in the populations and sampling methodology. We therefore considered it reasonable to combine the two datasets for our analyses. This created a more diverse sample, and from a practical point of view also boosted the number of cases reporting unilateral tinnitus.

### Variables for hearing asymmetry

A benchmark’ variable for asymmetric hearing was defined according to Jeffery et al. (2016) as an interaural difference of 20 dB or more in at least two consecutive frequencies at 0.5, 1, 2, 4 and 8 kHz. Four additional variables, which additionally quantify the *degree* of hearing asymmetry, were also calculated:

1. **MaxDiff:** the maximum mean interaural threshold difference of two adjacent frequencies (including thresholds at the frequency with the maximum interaural difference), spanning the range of thresholds at 0.5, 1, 2, 4 and 8 kHz as in Tsai, Sweetow et al. (2012).
2. **MaxDiffExt:** calculated as MaxDiff, spanning the range of thresholds at 0.125, 0.25, 0.5, 0.75, 1, 1.5, 2, 3, 4, 6 and 8 kHz, and including the mean difference from the available extended high frequencies (10, 12.5, and 14 kHz for the STOP dataset and 9, 10, 11.2, 12.5, and 14kHz for the BRC dataset). Thresholds at 0.75 and 1.5 kHz were not available for the STOP dataset and were calculated as the mean of the adjacent frequencies.
3. **AUCDiff:** the interaural difference of the area under the audiogram curve (integral) after logarithmically transforming frequencies to obtain equal distance per octave and interpolating in-between thresholds (including all available thresholds at 0.125-14 kHz).
4. **PTADiff:** the interaural difference of the mean threshold at 0.5, 1, 2, 4, and 8 kHz.

MaxDiff and MaxDiffExt emphasize the informational content of the two frequencies with the maximum interaural difference. In contrast, AUCDiff and PTADiff emphasize the overall average of the interaural difference. Another key difference is that MaxDiffExt and AUCDiff incorporate information from all available thresholds, whereas MaxDiff and PTADiff are limited to the mid-frequency octaves.

### Variables for tinnitus spatial perception

For both datasets the question ‘Where do you perceive your tinnitus?’ was asked, and response options were (a) in the right ear, (b) in the left ear, (c) in both ears equally, (d) in both ears but worse in the right or left ear, (e) inside the head, or (f) elsewhere (Langguth et al., 2007). Following Table 1, our variables for summarizing tinnitus spatial perception were:

1. (lateralized) unilateral
2. lateralized bilateral
3. non-lateralized bilateral

Throughout this report, the term ‘laterality’ is used to describe subgroups of unilateral and bilateral tinnitus, regardless of how the classification was done.

## Analysis

All analyses were conducted in R version 3.6.1 (R Core Team, 2019). R packages used included pROC (Robin et al., 2011), caret (Kuhn, 2015), glmnet (Friedman et al., 2010), missForest (Stekhoven, 2015, Stekhoven and Bühlmann, 2012), FSA (Ogle, 2017), and viridis (Garnier, 2018). Alpha level was set to 0.05 and for multiple comparisons p-values were adjusted using Holm’s method (Holm, 1979).

To address question 1, Receiver Operating Characteristic (ROC) curves were used to assess performance of hearing asymmetry variables for discriminating unilateral tinnitus (defined as the positive condition) from bilateral tinnitus (Robin et al., 2011). ROC curves are plots of the true positive rate (or sensitivity; proportion of correctly classified as positive of all positives) on the y-axis and the false positive rate (or 1 – specificity; proportion of wrongly classified as positive of all negatives) on the x-axis for different thresholds of a predictor. The area under the ROC curve (ROC AUC) takes values from 0 to 1, with 1 indicating excellent discrimination and 0.5 no discrimination capacity. The 95% confidence intervals for ROC AUCs were calculated using stratified bootstrapping (R package pROC; Robin et al., 2011). Delong’s method was used for comparison of ROC curves (DeLong et al., 1988), as implemented in the roc.test function from the pROC package (Robin et al., 2011). Results present the p-values for the pair-wise tests for statistically significant differences. Further, the ROC curve was used to define a cut off value to transform a numerical hearing asymmetry variable into a binary categorical variable. The best cut off was defined as the value that maximized the sum of sensitivity and specificity from the ROC curve (J-Index; Youden, 1950).

A further exploratory analysis compared performance of different operational definitions of binary categorical variables for hearing asymmetry in predicting tinnitus laterality. To do this, we calculated the specificity (proportion of being correctly classified as negative of all negatives), accuracy (fraction of all instances that are classified correctly), positive predictive value (proportions of being correctly classified as positive of all classified as positive), and negative predictive values (proportion of being correctly classified as negative of all classified as negative). Higher value for all these metrics indicates better performance.

To address question 2, box plots and frequency distributions were used to explore the relationship between hearing asymmetry and tinnitus spatial perception. Kruskal-Wallis test and post-hoc Dunn’s test were used to compare the distribution of hearing asymmetry across the different tinnitus spatial perception subgroups.

To address question 3, the associations between tinnitus laterality and various other phenotypic variables were assessed using Fisher’s exact tests and Wilcoxon tests. In addition, a multivariable logistic regression was used to assess the simultaneous effect of selected phenotypic variables in predicting tinnitus laterality. To avoid overfitting, the following protocol was applied for variable selection. First, a set of variables was selected by the authors. Then, univariable logistic regression models were fitted and the variables found significant were subsequently considered simultaneously into a multivariable logistic regression. The latter was fitted using least absolute shrinkage and selection operator (LASSO) (R package glmnet; Friedman et al., 2010). LASSO is a method for fitting linear models that includes a penalization for the sum of the absolute coefficients (Tibshirani, 1996). The method shrinks some coefficients to zero, allowing selection of the most relevant variables. Performance of the method was assessed using a 5-fold cross validation in an outer loop. The parameter lambda, which defines the penalty for the coefficients, was selected using 5 fold cross-validation in an inner loop (nested cross validation; see for example Varma and Simon, 2006), choosing the largest value for which error was within 1 standard error from the minimum (Breiman et al., 1984, Friedman et al., 2010). Cases with more than 20% missing values were excluded. Otherwise missing values were imputed using a random forest algorithm (R package missForest; Stekhoven, 2015, Stekhoven and Bühlmann, 2012).

## Results

### A hearing asymmetry emphasizing the maximum interaural difference across the full audiometric range most strongly discriminated unilateral from bilateral tinnitus

For the 571 cases reporting tinnitus, the four hearing asymmetry variables (MaxDiff, MaxDiffExt, AUCDiff and PTADiff) were compared to one another in their ability to predict tinnitus laterality. For each variable, the absolute values for hearing asymmetry were used as a marker for the degree of asymmetry. Only participants whereby tinnitus could be clearly discriminated as unilateral (left or right ears) or bilateral (both ears equally) (Table 1) were included in this analysis to avoid any difficulties in interpreting the findings which could be attributed to categorization bias.

For these data, ROC curves were plotted with each of the hearing asymmetry variables as predictors and tinnitus laterality as the outcome (Panel A, Figure 1), while Table 2 shows p-values from ROC AUC pairwise comparisons using DeLong’s test for correlated ROC curves. From visual inspection, differences between the ROC curves appeared to be marginal, and this was supported by the DeLong’s results which were mostly non-significant. A notable exception was that of the maxDiffExt metric which performed significantly better than AUCDiff in classifying tinnitus laterality (Table 2).

**Figure 1.**
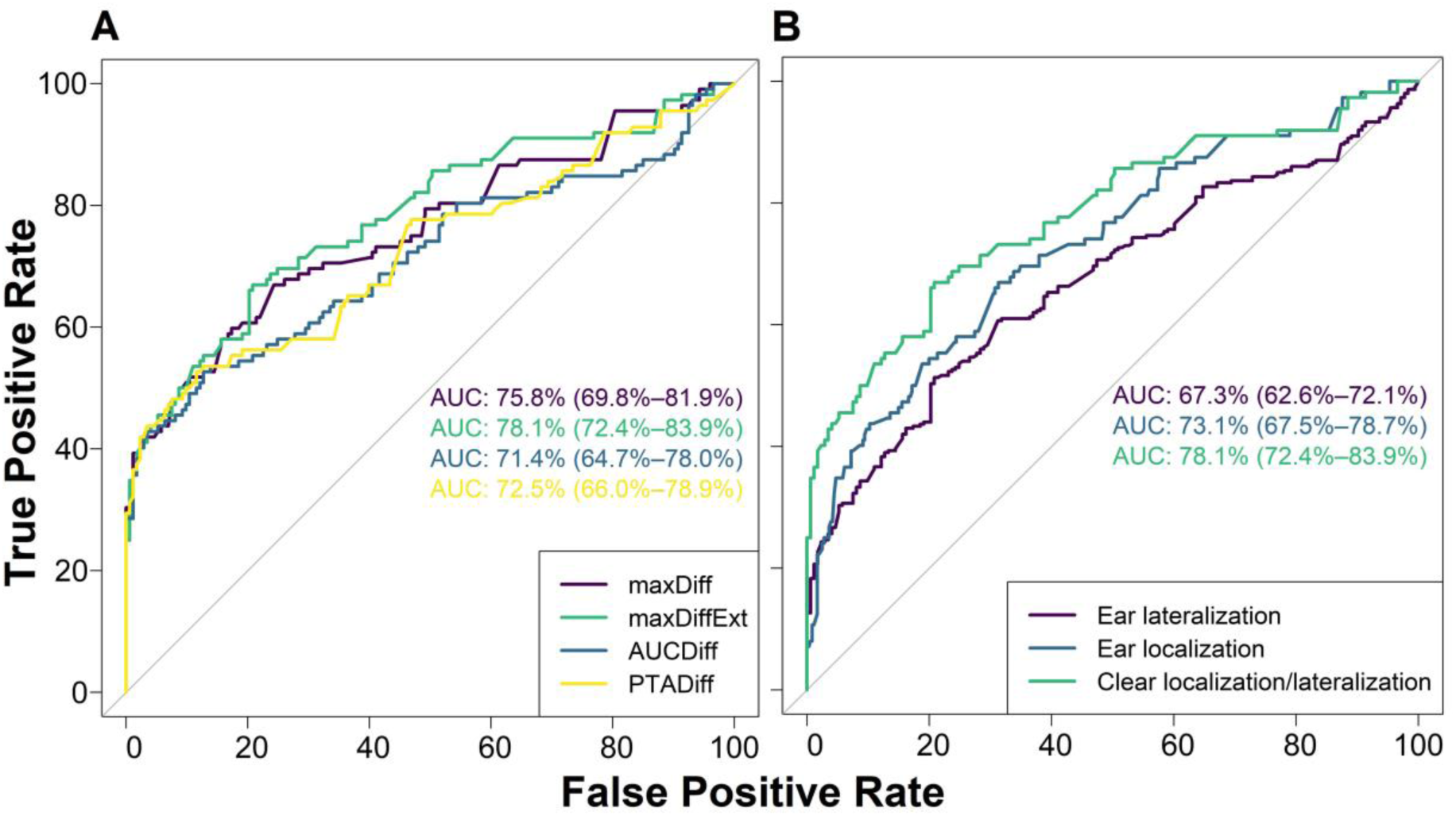
ROC Curves and AUCs using: A) the four hearing asymmetry variables (absolute values) as predictors and tinnitus laterality as outcome, and B) absolute MaxDiffExt as predictor and each of the different binary variables for tinnitus laterality as outcome. Unilateral tinnitus (versus bilateral) was coded as the positive outcome. MaxDiff: maximum interaural threshold difference mean of two adjacent frequencies including thresholds at 0.5-8 kHz; maxDiffExt: same as MaxDiff including thresholds at lower frequencies, half-octave frequencies and extended high frequencies; AUCDiff: interaural difference of the area under the audiometric curve including all available thresholds at 0.125–14 kHz; PTADiff: interaural difference of the mean threshold at 0.5–8 kHz; Ear lateralization: lateralized unilateral and bilateral versus non-lateralized bilateral; Ear localization: (lateralized) unilateral versus (lateralized and non-lateralized) bilateral; Clear localization/lateralization: (lateralized) unilateral versus non-lateralized bilateral.

**Table 2.** Pairwise comparison of AUCs of ROC curves 224 for the four hearing asymmetry variables.

|  | maxDiffExt | AUCDiff | PTADiff |
| --- | --- | --- | --- |
| maxDiff | 0.781 | 0.481 | 0.481 |
| maxDiffExt |  | 0.032 | 0.184 |
| AUCDiff |  |  | 0.781 |

P-values from Delong's test for correlated ROC curves (adjusted for multiple comparisons; Holm 1979).

We therefore conclude that the maxDiffExt metric was the preferred hearing asymmetry variable for subsequent subgrouping analyses. Not only did it perform best on the ROC evaluation, but also incorporated all available information obtained from the pure tone audiometry.

As used so far, the MaxDiffExt variable quantifies the degree of hearing asymmetry on an numerical scale. But for clinical decision making, a binary classification (akin to a ‘diagnosis’) is preferred as this clearly discriminates a person with symmetric hearing from a person with asymmetric hearing. The best cut off value for MaxDiffExt to define such a binary hearing asymmetry variable was found to be 14.54 dB (value that maximized sum of sensitivity and specificity). For practical purposes, 14.54 dB was rounded up to the nearest integer giving a recommended cut off of 15 dB. We therefore ascribed the label ‘symmetric hearing’ in all cases where the absolute maxDiffExt was <15 dB and ‘asymmetric hearing’ when the absolute maxDiffExt was ≥15 dB. This newly derived variable was called Asym15.

The performance of Asym15 in discriminating tinnitus laterality was compared to the performance of the Jeffery et al. (2016) benchmark. The latter showed high specificity and positive predictive value, but this contrasted with its rather poor sensitivity. Although Asym15 did not perform with the same specificity, it was a much more sensitive metric, performing better at correctly classifying positive cases (unilateral tinnitus) as true positive (Table 3).

**Table 3.**
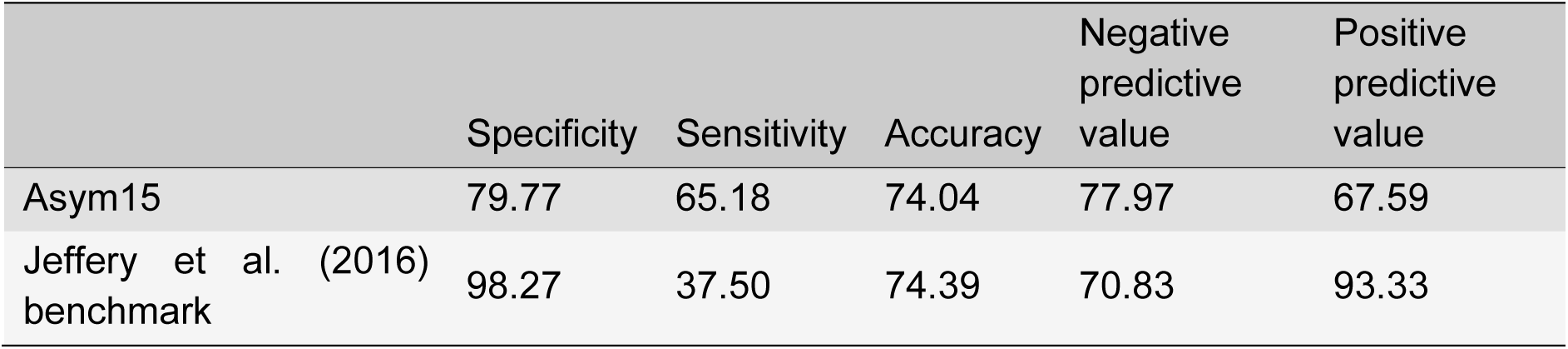
Performance of Asym15 and Jeffery et al. (2016) binary classification variables for hearing asymmetry.

In summary, for cases where the tinnitus spatial percept is unambiguous, we conclude that a hearing asymmetry variable emphasizing the maximum interaural difference across the full audiometric range appears able to most reliably discriminate unilateral from bilateral tinnitus.

### Merging lateralized bilateral tinnitus with unilateral tinnitus weakened the association with hearing asymmetry

Our analysis so far excluded cases where the laterality of the tinnitus spatial percept was somewhat ambiguous (i.e. cases of lateralized bilateral tinnitus in both ears, but worse on one side). But since these cases represent 32.9% (188/571) of the full tinnitus dataset, they should preferably not be ignored. A follow-on analysis was therefore conducted to investigate the effect of adding these participants into the ROC computation. The exploratory question that we asked was how would adding these participants affect the good performance of the maxDiffExt in discriminating unilateral from bilateral tinnitus?

The benchmark was the previous dataset comprising only participants whereby tinnitus could be clearly discriminated as unilateral (left or right ears) or bilateral (both ears equally). This condition is termed ‘clear ear localization/lateralization’. Two comparator datasets were created. One discriminated unilateral (tinnitus in left or right ears) from bilateral (tinnitus in both ears equally, plus tinnitus in both ears but worse on one side). This condition is termed ‘ear localization’. Another discriminated lateralized (tinnitus in left or right ears, plus tinnitus in both ears but worse on one side) from non-lateralized (tinnitus in both ears equally). This condition is termed ‘ear lateralization’.

ROC curves were plotted with the maxDiffExt as the predictor and each of the three different conditions defining tinnitus laterality as the outcome (Panel B, Figure 1), while Table 4 shows p-values from ROC AUC comparisons using DeLong’s test for uncorrelated ROC curves. From visual inspection, differences between the ROC curves appeared to be marginal, but notably the DeLong’s results indicated that ear lateralization performed significantly worse than the benchmark condition in classifying tinnitus laterality. Ear localization did not significantly differ from the benchmark condition.

**Table 4.** Pairwise comparison of AUCs of ROC curves for the different binary variables for tinnitus spatial perception.

|  | Ear localization | Clear localization/lateralization |
| --- | --- | --- |
| Ear lateralization | 0.2534 | 0.014 |
| Ear localization |  | 0.253 |

P-values from Delong's test for uncorrelated ROC curves (adjusted for multiple comparisons).

We therefore conclude that one should not consider participants who report their tinnitus in both ears but worse on one side, as being equivalent to participants who report a unilateral tinnitus clearly in the left or right ear. Doing so reduced the discriminative power of hearing asymmetry for tinnitus laterality subgroups.

### Association between asymmetric hearing and a unilateral tinnitus reported on the side of the worse hearing ear

Question 2 addressed how the pattern of hearing asymmetry differed between tinnitus and non-tinnitus cases, and for those reporting tinnitus, across different spatial tinnitus percepts. The MaxDiffExt data computed for all participants in the full dataset (n=833) were displayed using box plots (Figure 2); data points falling between the dashed lines indicate symmetric hearing (Asym15). On visual inspection, there was a trend towards an association between asymmetric hearing and unilateral tinnitus on the side of the worse ear. Nevertheless, many unilateral tinnitus cases had symmetric hearing. The remaining tinnitus cases all showed a similar pattern to one another, tending towards symmetric hearing. The same was also true for the non-tinnitus cases, albeit with some extreme deviations. Distributions of hearing asymmetry differed across different tinnitus spatial percepts (Kruskal-Wallis chi-squared = 84, degrees of freedom = 5, p-value < 0.001). The Dunn post-hoc tests showed that lateralized bilateral tinnitus was heterogeneous, with ‘both, more left’ tinnitus being significantly different to ‘both, more right’ tinnitus (Supplementary Table 5).

**Figure 2.**
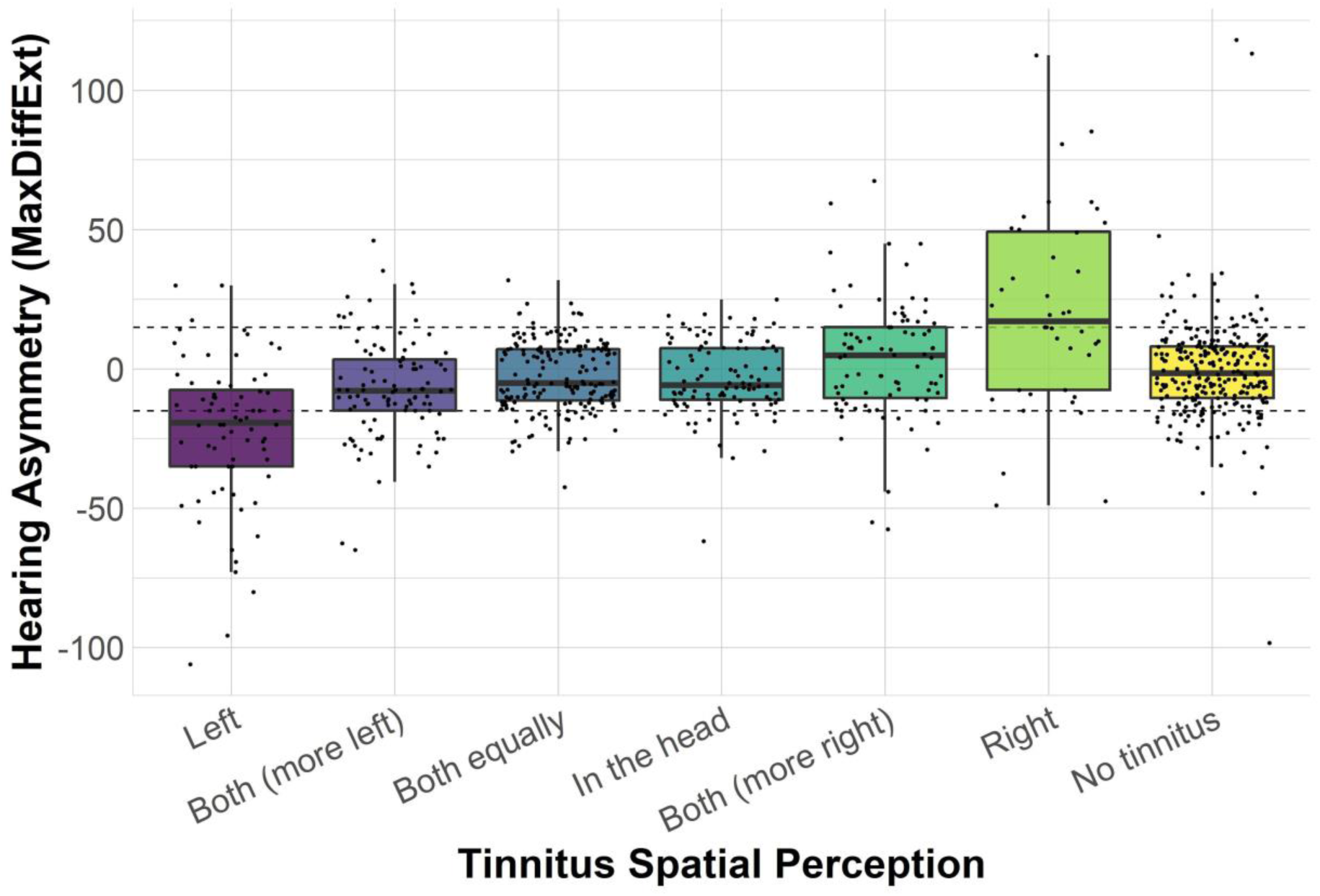
Box plots of MaxDiffExt (right minus left thresholds) for tinnitus cases reporting different tinnitus spatial perceptions, and for the non-tinnitus cases. The dashed line shows the 15 dB asymmetry threshold, defining Asym15.

Data were displayed in an alternative format by using Asym15 to categorize individuals into symmetric or asymmetric hearing (Figure 3). The majority of participants (67.3%) in the full dataset had symmetric hearing. The non-tinnitus group and the group reporting a non-lateralized tinnitus (both ears equally or in the head) had the highest proportion of symmetric hearing. Many of these had clinically normal hearing (no threshold higher than 20 dB, Supplementary Figure 1). Asymmetric hearing was present in 35.9% of tinnitus cases and 25.6% of non-tinnitus cases. The unilateral tinnitus group had the highest percentage of asymmetric hearing (58.0% with ipsilateral asymmetric hearing). This frequency distribution confirmed the association between asymmetric hearing and unilateral tinnitus on the side of the worse ear. Nevertheless, there were also many cases with unilateral tinnitus and symmetric hearing (34.8%). Notably, there were some cases with contralateral hearing asymmetry in the lateralized bilateral tinnitus (13.3%) and the unilateral tinnitus (7.1%) subgroups (Figure 3).

**Figure 3.**
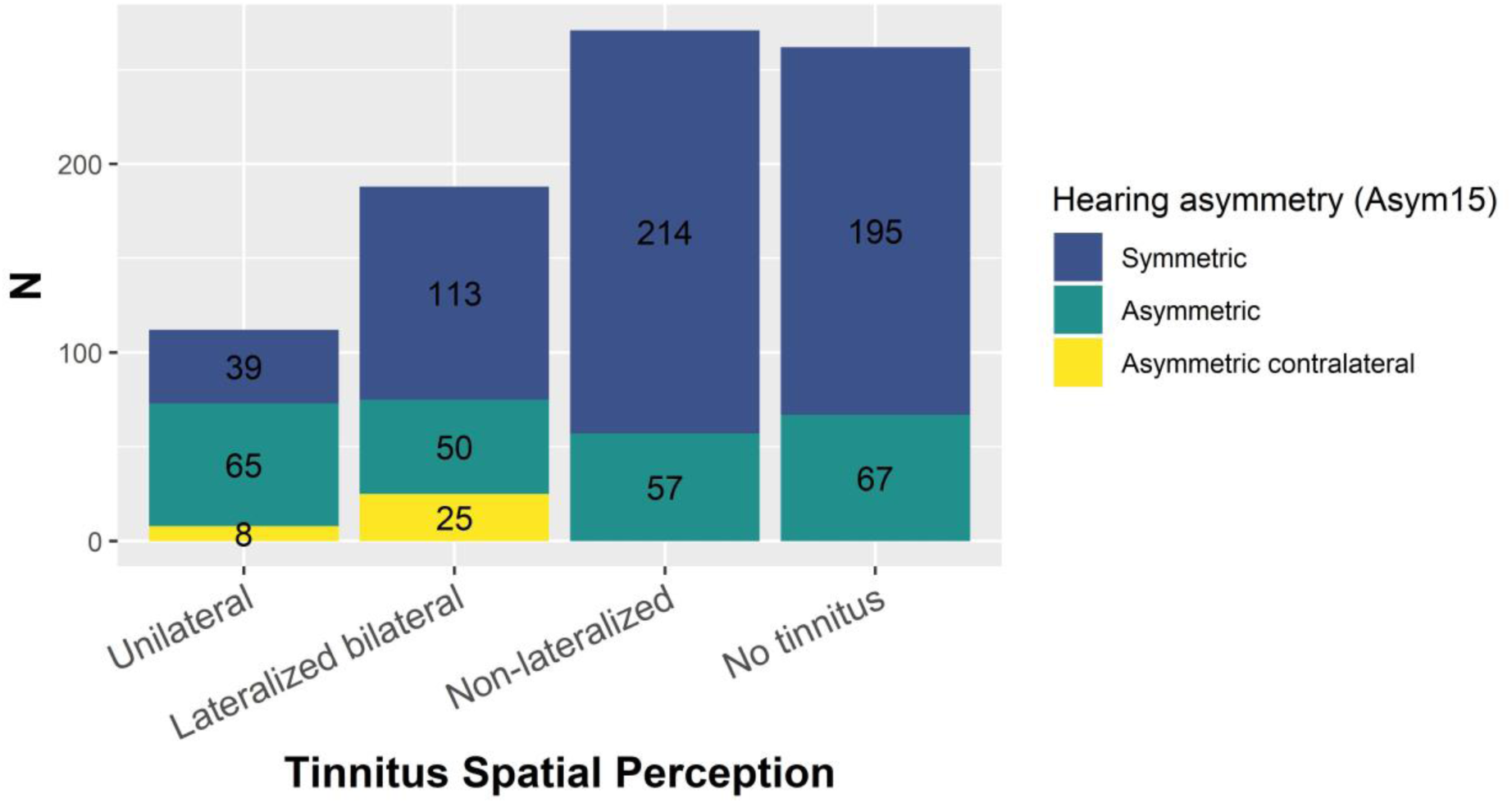
Frequency of symmetric and asymmetric hearing for unilateral, lateralized bilateral, and non-lateralized tinnitus, and non-tinnitus cases. Contralateral asymmetry is presented separately for lateralized cases.

In summary, we observed a trend towards an association between hearing asymmetry and tinnitus spatial perception; specifically between asymmetric hearing and a unilateral tinnitus reported on the side of the worse hearing ear. This indicates a potential criterion for subgrouping people with tinnitus.

### Spatial tinnitus perception is an important variable for tinnitus phenotyping

Question 3 compared unilateral (left or right ears) and bilateral (both ears equally) tinnitus on a number of phenotypic variables, investigating whether any of these might be informative for predicting tinnitus laterality. Compared to bilateral tinnitus, participants with unilateral tinnitus were older, with older age at tinnitus onset, and shorter tinnitus duration (Table 5). In addition, they had higher hearing asymmetry, more often used a hearing aid unilaterally, and were annoyed by tinnitus for a higher percentage of time. The multivariable LASSO logistic regression model identified hearing asymmetry, hearing aid use, and age at tinnitus onset as predictors of tinnitus laterality. The 5-fold cross-validated ROC AUC of the regression method was 84.2%, indicating very good predictive power.

**Table 5.** Comparison of unilateral (left or right ears) and bilateral (both ears equally) tinnitus.

|  | All | Unilateral | Bilateral | Statistics |
| --- | --- | --- | --- | --- |
| All | 285 | 112 | 173 | - |
| *General individual characteristics* |  |  |  |  |
| Age (y) | 54.7, 13.56,<br>n=284 | 58.85, 12.61,<br>n=111 | 52.03, 13.51,<br>n=173 | W=6991,<br>p=0.001<sup>z</sup> |
| Gender (male/female) | 131/154 | 55/57 | 76/97 | p=1 |
| Handedness<br>(both/left/right) | 4/23/257 | 1/9/101 | 3/14/156 | p=1 |
| *Hearing function and other comorbidities* |  |  |  |  |
| Absolute maxDiffExt (dB) | 17.8, 17.74,<br>n=285 | 28.63, 23.22,<br>n=112 | 10.79, 6.77,<br>n=173 | W=4235.5,<br>p<0.001* |
| Hearing aid use (both<br>sides/unilateral/none) | 19/14/221 | 6/14/76 | 13/0/145 | p=0.006* |
| TMJ disorder (no/yes) | 236/32 | 91/17 | 145/15 | p=1 |
| Balance disorder (no/yes) | 188/82 | 70/38 | 118/44 | p=1 |
| Headaches (no/yes) | 203/70 | 76/32 | 127/38 | p=1 |
| HQ score (0-42) | 15.07, 8.2,<br>n=279 | 14.32, 7.77,<br>n=106 | 15.53, 8.44,<br>n=173 | W=9778,<br>p=1 |
| *Tinnitus-related characteristics* |  |  |  |  |
| Age at tinnitus onset (y) | 39.08, 17.7,<br>n=206 | 45.87, 16.13,<br>n=94 | 33.38, 16.99,<br>n=112 | W=3155.5,<br>p<0.001* |
| Tinnitus duration (y) | 14.98, 13.09,<br>n=207 | 12.38, 12.66,<br>n=95 | 17.17, 13.11,<br>n=112 | W=6759.5,<br>p=0.009<sup>z</sup> |
| Tinnitus annoyance (%) | 24.43, 25.83,<br>n=278 | 29.27, 26.4,<br>n=110 | 21.26, 25.02,<br>n=168 | W=7247,<br>p=0.022<sup>z</sup> |
| Tinnitus loudness rating<br>(0-100) | 42.9, 22.7,<br>n=274 | 43.22, 20.24,<br>n=109 | 42.69, 24.24,<br>n=165 | W=8619,<br>p=1 |
| Pulsatile tinnitus (no/yes) | 258/21 | 104/6 | 154/15 | p=1 |
| Tinnitus influenced by<br>stress (no<br>effect/reduces/worsens) | 103/57/118 | 44/17/48 | 59/40/70 | p=0.244 |

Table presents frequencies for categorical variables and mean, standard deviation and sample size for numerical variables. Statistical tests: Fisher's exact tests for categorical variables and Wilcoxon tests for numerical variables. P-values are adjusted for multiple comparisons.
<sup>z</sup>Significant coefficient in simple regression and zero coefficient in LASSO regression; \*Significant coefficient in simple regression and non-zero coefficient in LASSO regression; HQ: Hyperacusis Questionnaire; TMJ: Temporomandibular joint.

In summary, unilateral and bilateral tinnitus groups differed in a number of statistically significant ways. The modelling work confirmed a relationship between hearing asymmetry and tinnitus spatial perception and suggested that spatial tinnitus perception may be informative as a criterion for subgrouping people with tinnitus.

## Discussion

The principle findings of this study were:

1. A hearing asymmetry variable emphasizing the maximum interaural difference across the full audiometric range most reliably discriminated unilateral and bilateral tinnitus. Grouping lateralized bilateral tinnitus with unilateral tinnitus weakened this discrimination.
2. There was an association between asymmetric hearing and a unilateral tinnitus reported on the side of the worse hearing ear.
3. Unilateral and bilateral tinnitus were phenotypically different.

The strength of the study is in using data drawn from two distinct sampling populations (i.e. from people participating in tinnitus clinical trials and from people with tinnitus recruited from a population-based cohort) and two countries (i.e. UK and Sweden). Combining the two datasets for our study led to a large and diverse sample that allowed us to statistically explore tinnitus heterogeneity focusing on the relationship between tinnitus spatial perception and hearing asymmetry. We expect that the large and diverse sample would make our findings generalizable to other datasets, and we greatly encourage attempts of replication.

### Hearing asymmetry and tinnitus spatial perception

Examining different variables for hearing asymmetry, there was a similar performance in discriminating tinnitus laterality. Nevertheless, a variable emphasizing the maximum interaural difference (mean difference of two adjacent frequencies), using all available thresholds, demonstrated the best performance. The optimum threshold for asymmetric hearing was 15 dB. This finding is in agreement with Tsai et al. (2012) who also investigated how different hearing asymmetry variables associated with tinnitus spatial perception. Specificity, sensitivity, and positive predictive value were 80, 65 and 68% respectively, as compared to 71, 59, and 76% in Tsai et al. (2012). The higher specificity in our study could be due to the exclusion of the non-lateralized bilateral cases and the additional frequencies used for calculation of the asymmetry variable.

Regarding the ambiguous cases in which tinnitus is reported in both ears but greater on one side, to our knowledge, only one previous study has reported their hearing asymmetry profile, presenting only the mean thresholds for each ear per tinnitus subgroup (Nuttall et al., 2004). In our study, hearing asymmetry for individual cases and frequency of symmetric and asymmetric hearing were assessed. We showed that lateralized bilateral cases represent a large proportion of the tinnitus population and, although the majority had symmetric hearing, asymmetric hearing was common. This should be considered in future studies when deciding to group this type of tinnitus with either unilateral or non-lateralized bilateral tinnitus.

It is not clear why for some tinnitus cases hearing asymmetry is not predictive of tinnitus laterality. One possibility is that pure tone audiometry at specific frequencies is not enough, and that more detailed hearing assessment would reveal hearing loss corresponding to the spatial perception of tinnitus (Xiong et al., 2019). One recent study analyzed characteristics of 62 unilateral tinnitus cases with better mean hearing threshold on the tinnitus side (Lee et al., 2019). About one fourth of these cases were shown to be associated with fluctuating hearing loss and in seven cases there were indications of somatic tinnitus.

### Tinnitus laterality subgroups

When we examined phenotypical characteristics differentiating unilateral from bilateral tinnitus, the most robust differences were in hearing asymmetry, hearing aid use, and age at tinnitus onset. In addition to these, subgroups differed in age, tinnitus duration, and percentage of time being annoyed by tinnitus. At least seven other studies with sample sizes larger than 50 have compared characteristics of unilateral and bilateral tinnitus (Gabr, 2011, Hallam et al., 1984, Koning and Koning, 2018, Pan et al., 2009, Vanneste et al., 2011, Yang et al., 2015, Zagólski and Stręk, 2017). Interestingly, none of these reported hearing asymmetry across groups.

Comparing our results with other studies, a common finding is that unilateral tinnitus corresponds to shorter tinnitus duration than bilateral tinnitus (Pan et al., 2009, Zagólski and Stręk, 2017). One interpretation is that unilateral tinnitus might evolve to bilateral tinnitus with time (Pan et al., 2009). In our study, unilateral tinnitus was also characterized by older age at tinnitus onset. This is in agreement with the findings of Maas et al. (2017), who showed that in a twin cohort heritability was much higher for bilateral tinnitus (0.56) than unilateral tinnitus (0.27). Considering this, another potential explanation for the difference in tinnitus duration is the earlier onset of the more genetically influenced bilateral tinnitus. In addition, bilateral tinnitus was shown to be associated with a higher percentage of prolonged exposure to excessive noise than unilateral tinnitus (Zagólski and Stręk, 2017), suggesting that a combination of genetic and environmental factors might trigger an earlier onset.

Yang et al. (2015) found that bilateral tinnitus cases were older with a higher tinnitus burden. In contrast, in our study, as in Zagólski and Stręk (2017), unilateral cases were older. With regards to tinnitus impact, unilateral cases in our dataset were annoyed by their tinnitus for a greater percentage of time. A higher burden of tinnitus for the unilateral tinnitus cases was also found by Song et al. (2018). The discrepancies with the findings from Yang et al. (2015) could be due to differences in the sampling population characteristics. For example, in Yang et al. (2015) there was a high percentage of normal hearing, especially for unilateral tinnitus (63.8%). In our dataset, only a few tinnitus cases had normal hearing and these were mainly non-lateralized tinnitus cases (Supplementary Figure 1).

Overall, there is evidence suggesting that subgroups of tinnitus with different spatial perception might be associated with different underlying mechanisms. Tinnitus spatial perception is associated with hearing asymmetry, but further research is needed to understand why hearing asymmetry is not always predictive of tinnitus laterality. In addition, unilateral tinnitus compared to bilateral, seems to have an earlier onset age and has been repeatedly shown to have a shorter duration than bilateral tinnitus. This evidence supports the recommendation that tinnitus spatial perception should be used to define phenotypically more homogeneous tinnitus subgroups for tinnitus research and clinical practice.

### Limitations and future considerations

The main limitation of our study is that, although the sample size was relatively large compared to previous studies, it is still small considering the high dimensionality of tinnitus. In addition, our combined dataset did not include some potentially important variables, such as family history of tinnitus or self-reported tinnitus severity, because they were collected using different measures across the two datasets. Pure tone audiometry methodology was also different in the STOP and BRC datasets. We do not expect this to influence our results as automated audiometry has been shown to be comparable to manual methods (Mahomed et al., 2013), and any systematic difference would be eliminated in the asymmetry indices because these reflect a difference between two measurements. Nevertheless, we refrained from comparing overall hearing thresholds across unilateral and bilateral subgroups, because participants in each subgroup were not balanced across the STOP and BRC datasets. Other information missing from our datasets that would be important for characterizing subgroups of tinnitus is brain imaging and genetic profiling.

Previous efforts to standardize tinnitus research has allowed us to combine independent datasets for this analysis (Langguth et al., 2007). Such efforts should be reinforced to allow the creation of even larger datasets with a broader spectrum of information per participant, to further understand tinnitus heterogeneity (Schlee et al., 2018).

## Data Availability

Please contact the corresponding author [EG] for queries about accessing the data that support the findings of this study. Restrictions apply to the availability of these data, which were used under license for this study.

## Acknowledgments

Thanks to Eirini Genitsaridi for the valuable discussions on data analysis.

